# The impact of the fragile health system on the implementation of health policies in Yemen

**DOI:** 10.1101/2020.09.25.20200741

**Authors:** Taha Hussein, Fekri Dureab, Raof Al-Waziza, Hanan Noman, Lisa Hennig, Albrecht Jahn

## Abstract

**Background:** The on-going humanitarian crisis in Yemen is one of the worst in the world, with more than14 million people in acute need. The conflict in Yemen deteriorated the country’s already fragile health system and lead to the collapse of more than half of the health facilities. Health system fragmentation is also a problem in Yemen, which is complicated by the existence of two health ministries with different strategies. The aim of this study is to evaluate the effect of health system fragmentation on the implementation of health policies in Yemen across the global agendas of Universal Health Coverage (UHC), Health Security (GHS) and Health Promotion (HP) in the context of WHO’s priorities achieving universal health coverage, addressing health emergencies and promoting healthier populations.

**Methods:** The study is qualitative research using key informant in-depth interviews and documents analysis.

**Results:** There are many health stakeholders in Yemen, including the public, private, and NGO sectors - each with different priorities and interests, which did not always align with national policies and strategies. The WHO and Ministry of Public Health and Population (MoPHP) are the main supporters to implement all policies related to the UHC, GHS and HP agendas. Interestingly, initiatives initially pursuing a health security approach to control the cholera epidemic realigned with the UHC concept and moved from an initial focus on health security, to propose a minimum health service package, a classical UHC intervention. Overall, Universal Health Coverage is the most adapted agenda, health security agendas were highly disrupted due to conflicts and health staffs were caught unprepared for emerging outbreaks. The health promotion agenda was largely ignored.

**Conclusion:** Restoring peace, building on synergies between the three health agendas through joint planning between the MoPHP and other health actors are highly recommended.

## Introduction

Yemen is the most impoverished country in the Middle East. Over half of the population lives below the poverty line with only limited access to essential health services ^1^. The on-going humanitarian crisis in Yemen is one of the worst in the world, with more than 14 million people in acute need ^2^. Poverty, political instability, and civil war destroyed Yemen’s infrastructure, collapsed the economy, displaced the population and resulted in the breakdown of public institutions in the country. Around two million children under five are malnourished and over one million pregnant and lactating women facing imminent death from malnutrition or maternal complications if urgent care is not provided ^3^.

Since March 2015, the Yemen armed conflict destroyed the national infrastructure, including the health sector. As a result, almost 50% of Yemeni health facilities are not functional, and those health facilities which continue to function lack specialists, equipment and medicines. ^3^. The health system in Yemen became more fragmented due to the prolonged armed conflict and its component’s previous fragility. The health system is further challenged by fragmentation in oversight between two separate ministries of health and the reduction of the national health budget. Out of pocket (OOP) payments have reached an unprecedented level, at over 80%, forcing the Yemeni population to face impoverishment in order to access life-saving health care ^4,5^.

With its basis in the principles of Health for All, achieving universal health coverage (UHC) for all people is a predominant global goal of the SDGs, although it is vital that each country considers their individual development of health policies and strategies over time to achieve it ^6^. Reaching this ultimate goal of UHC, the efforts of communities, national governments, and international agencies, should be integrated to prevent fragmentation within the health system and deliver high-quality health care while avoiding catastrophic impoverishment of poor populations as a result of seeking health care ^7^.

The health system in Yemen suffers from multiple problems including, but not limited to, the lack of financial resources, essential medicine, and qualified health staff, which affects the delivery of the essential health care services ^8^. Most of the physicians are concentrated in the urban areas although most of the population live in rural areas ^8^. Only about one-third of the rural population is covered by health services ^8^.

The aim of this study is to evaluate the effect of health system fragmentation and forgone synergies on the implementation of national health policies in Yemen. The study is particularly concerned with the fragmentation among the triangle of the three leading global health agendas: universal health coverage (UHC), global health security (GHS), and health promotion (HP). The study is also interested in the coordination mechanisms amongst different health sector actors, their synergistic links, and how these affect the agenda-setting on the national scale. The study was inspired by the Lancet Commission on synergies between UHC, GHS, and HP that aims to overcome fragmentation and realize the potential for coherence in global health ^9^. The WHO’s global programme of work 13 highlights the importance of a synergistic approach to these agendas ^10^.

## Methods

### Study Design

This study uses qualitative research grounded on case study design. This approach is imperative to explore participants’ perceptions and thoughts on Yemeni health system fragmentation. The study utilizes the synergies between UHC, GHS and HP from WHO’s GPW 13 (2019-2023) as an integrative framework to understand the current state of health system fragmentation in Yemen ^9^. The synergistic direction of this framework is key to understand the interplay of these three major global health priorities and essential for Yemen to identify insights and recommendations.

### Sampling

The researcher employed purposive sampling to select key health informants to be interviewed. Key informants were selected purposefully from the outcomes of the mapping exercise. Twenty key informants were invited to participate via email. The investigator has chosen the key informants equally from both health ministries that were formed during the conflict in the country and it was part of this fragmentation.

### Data Collection

This study takes a qualitative design by mixing two methods, the document analysis and in-depth interviews with key informants for the main health stakeholders in Yemen.

#### A: Document Analysis

The document analysis is an appropriate data collection method for this study as resources about Yemen are fairly limited and the health agendas, at the core of this study, are relatively new to the country. Twenty-two documents and reports related to Yemen’s health system were selected based on the following keywords:

“National health policy”, “Universal health coverage”, “Health service delivery”, “Health security”, “Health promotion”, “Fragmentation”, “National strategies”, “Fragile health system”, “Yemen profile”, “Yemen DHS”, “Minimum service package”, “Health stakeholders”, and “Ministry of Health Yemen”.

The document analysis alone did not provide enough information to answer the study questions and their main objectives. The gaps that are produced from the deficiency of clear answers in the document analysis was filled from the information conducted from the IDIs of the key informants and experts from the field.

#### B: Key Informant Interview

Overall, twenty key informants were contacted via E-mail to invite them to participate in this study as a key informant. Choosing the key informants was dependent on key characteristics: (1) their position in the MoPHP and the NGOs and, (2) their experience in the health system structure and the national health policies in Yemen. Out of twenty invited participants, six invitees refused to participate, and the other three did not replay for our e-mails. Eleven participants confirmed their participation via email, hence the final sample size for this study. Among the Eleven participants, 5 of them are from UN agencies and other NGOs, 3 are from the MoPHP of Yemen and the remaining 3 are independent consultants’ expert in the health system of Yemen.

The interviewees were conducted in English and Arabic languages. Due to the bad situation and current conflict in Yemen, the interviews were conducted via phone or video call (Skype). The investigator utilized an in-depth interview guide and took response notes for the participant during interviews. These audio recording and notes were securely saved.

### Reflexivity

The investigator acknowledges that his 11 years of working experience in both government and NGOs in Yemen may have influenced the way he sees the data and the way he pursued data analysis. Considering the importance of self-reflection at all stages, the investigator kept notes on his preconceptions and his past experiences and struggles in the health system. The constant dialogues with supervisors and self-reflection of the investigator may have ensured transparency of the research process. To ensure the credibility of study results, the supervisors checked the developed preliminary codes and themes.

### Data Analysis

All the eight interviews were transcribed verbatim in English language by the investigator. The investigator gave a special secure number for each participant (from one to eight) for anonymity purposes.

Interview transcripts were analysed manually using qualitative content analysis ^11^. Four data analysis steps were conducted to gain insight into the key informants’ perception of health system fragmentation. First, the investigator started the interpretation of the data by reading each interview transcripts and underlining striking statements. Second, all underlined statements were coded across each interview undergoing inductive analysis. Third, all codes were grouped into two themes: positive perceptions and negative perceptions. Finally, the investigator re-reads all statements in both themes to reflect on the overarching health worker’s perception about health system fragmentation. After the investigator reviewed all the documents retrieved and integrate it into the major themes – these documents complemented the study results. The review of governmental documents, stakeholder’s mapping and exhaustive literature review provided triangulation of the data. Collaborative discussion with the supervisors about the themes and findings was conducted for appropriate recommendations. After the integration of IDIs and documents, an exhaustive literature review was conducted to synthesize and ground the evolved findings.

### Ethics

The study is in line with the Declaration of Helsinki looking at getting ethical clearance, informed consent, ensure privacy and confidentiality of the research participants, and also covering the risk and the benefits of the research to the participants ^12^. Ethical clearance to conduct the study was sought from the Ethical Committee of the Ruprecht-Karls-Universität Heidelberg. Individual verbal and written informed consent was also sought from the participants.

## Results

### 1. Stakeholder Analysis

Figure **1** shows the potential stakeholders in Yemen, there are a few differences in stakeholders between the north and south of Yemen. Yemen has two different governments, one in the south and the other in the north. However, in the northern areas, a new powerful institution was created in 2017 called The National Authority for the Management and Coordination of Humanitarian Affairs (NAMCHA) as a supreme authority to manage and coordinate all humanitarian interventions. The local NGOs in the southern part of the country are not able to work in the northern part and vice versa, although there is an exception in a few cases, while the international organizations are working in both parts of the country.

**Figure 11:**
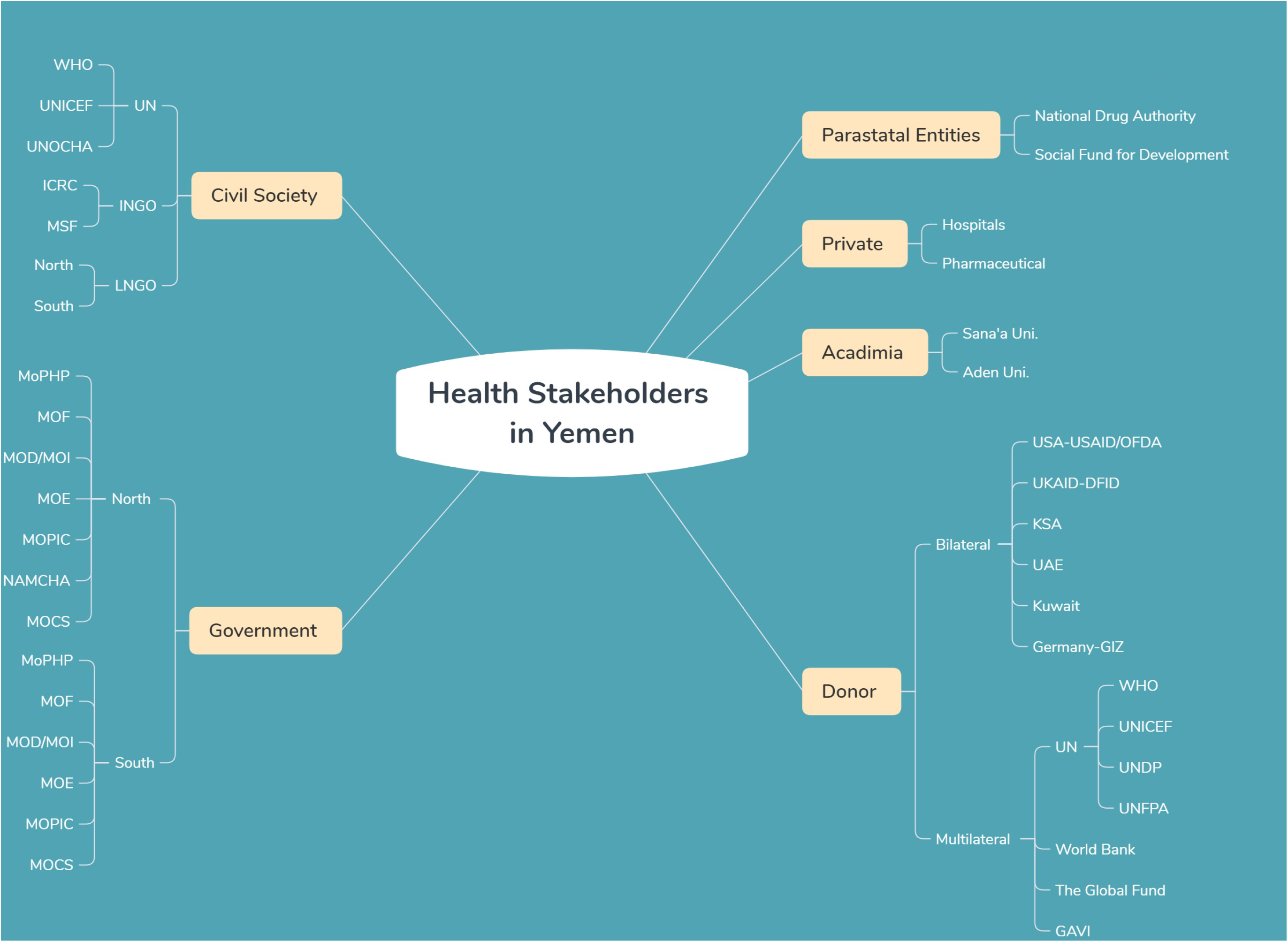
The potential stakeholders in support of the three agendas in Yemen

Majority of the key informants recognized the role of stakeholders in the development of national health policies and the MoPHP is considered the main stakeholder interested in implementing these strategies. The WHO has the same interest in the national and global level. Some key informants cannot separate the UHC and HP agenda. The MoPHP, WHO and other organizations prioritized UHC and HP in their agendas, while the health security was prioritized by other international donors like the US. This is considered as a negative influence on the health system, the implementing agencies were asked to label their projects as an emergency or as responding to global threats across the borders, and they may use this health security agenda as an excuse or justification to disperse the funds ^13,14^.

> “*We will speak about health system represented by the MoPHP, this a major stakeholder, and what we call them the governorate health offices […], then you could speak about the parastatal entities, and I would mention here very quickly the National Drug Authority, is a major stakeholder, which was always involved in these processes […], there was also the Social Fund for Development as being one of the major stakeholders who had some much investment in the health sector, you speak also about the line ministries, the ministry of planning and international cooperation (MOPIC), the ministry of finance (MOF), the ministry of civil services (MOCS), and then you have the donor community and what we call it development partners, at that moment of time there were 4×4, the four UN agencies; speaking about the WHO, UNICEF, UNFPA, and World Bank and the other four [are] developmental partners; GIZ, European Commission, USAID and DIFD*” **Informant # 6 (MoPHP)**.

Local NGOs were recognized by few key informants as effective partners during the crisis in Yemen. The reasons being easy and rapid access to the local communities and less costly in comparison to international NGOs.

> *“Local NGOs, there are some studies show that the local actor is most effective than global partners, we are talking about cost-effectiveness, so local areas (district or sub-district) they are the first to react, they are cost-effective, […*.*], they are there, they do not wait, they do not have this long arm of information and response, if we want to do that through UNICEF or WHO it will cost 100 times more and it will come 10 days later, so if you want to reduce the cost, we have to be open to the local actors”* Informant # 3 (Int. Consultant in Yemen).

### 2. Implementing the global health agendas in humanitarian context

The main focus of activity of the many policy actors was related to the mandate of their organization, however, a clear shift from development interventions toward humanitarian response was noticed in most of the actor’s agendas. The WHO and MoPHP are the main supporters to implement all policies related to the UHC, GHS and HP agendas. The Yemeni health system is extremely dependent on donors; the collapse of economic stability and lack of domestic budget for health during the Yemen crisis forced MoPHP in a weak position while trying to pursue the implementation of the National Health Policy (NHS). The power of the Ministry of Finance (MOF) was also minimized due to the crisis. The World Bank and UNICEF have also a strong influence in implementing the current policies regarding increasing access to health services and controlling outbreaks. Some national institutions are not viewed as supportive and instead blocked the implementation of several interventions in the field.

Moreover, participants mentioned that international NGOs adapt themselves according to the current conflict situation and the flow of funds in the country. They change their mechanism of work focusing more on emergency and health security rather than development and recovery.

> *“Maybe I will talk about NGOs, they adapted by changing the way they work, now there is a tendency to work in emergency and preparedness and health security, so if you noticed the projects that were implemented in the last three years or since the emergence of the conflict, you can see that there is change in the program design, […], all of these were based on cholera response mainly and other communicable diseases, so that was the change that happened, it was negative in the fact that they are seeking funding so, in order to get funds you have to abide by these roles and regulations, and by changing the nature of their work, it is also justified since we have in active conflict. However, we still have many areas within Yemen that did not have a conflict, or they do not have an active conflict for the time being, so these areas should have different strategies or a different approach to implement health response”* **Informant # 2 (Local NGO)**.

There are many factors that affecting the implementation of the three global health agendas. Key informants reported that there are some stakeholders who have an influence on the implementation of these strategies; and they put challenges and barriers in front of the humanitarian organizations, because the policymakers in the MoPHP are not aware of the importance of the prioritizing actions in the community.

> *“If I think about the negative influence, it’s difficult because sometimes, people, they do not mention it, but their action can be negative (intentional or unintentional). For example, if you see in the north of Yemen having a new body which is parallel to the Ministry of Planning and International Cooperation (MOPIC) called NAMCHA, which is really hindered many activities or put challenges in front of the humanitarian organization****” Informant # 1 (National Consultant)***.
>
> *“For the local authorities I think the role is very limited, maybe the authorities in the north are more powerful or more influence the de facto authorities compare to the counterpart in the south, and the influence mainly on channelling the fund or directing the fund, they have more powerful say in the north”* ***Informant # 2* (Local NGO)**.

Some key informants revealed that the implementation of these agendas is affected by the lack of awareness regarding the importance of these agendas within medical students and among health staff at central and peripheral levels of MoPHP including the decision-makers. Furthermore, the role of the UN is very weak to promote the strategies at the national level.

> “*We have conducted a small study on the knowledge and awareness of the primary health care, UHC and HP targeting students of the health science and national health staff and leaders at the MoPHP. We found that 80 – 90% of the medical students had no or limited knowledge about these agendas. If we ask people from the higher to lower levels of MoPHP, they may hear about these agenda, but did not implement and reflect them into indicators to monitor the progress of their achievement by 2030 on the ground. Certainly, as a decision-maker, he has health targets to achieve but without awareness that may serve these strategies at central or government levels of MoPHP. From my experience, even in the big UN organizations at the local level, they do not talk about these strategies with MoPHP”* **Informant # 4 (MoPHP)**.

One of our participants reported on the HP agenda implementation in Yemen in which UNICEF supports this agenda through the communication for development program. The program focuses on the implementation of the HP at the community levels, for instance by promoting immunization campaigns and increasing the hygiene awareness on prevention of infectious diseases through engagement in the community and by coordination with other sectors.

> “In *UNICEF for example, there is a section called C4D (Communication For Development), and [in] this section we are focusing on the community and how to promote health among the community, so we are engaging with the community themselves, either host communities, IDPs, and that’s happening in each activity that we are doing, especially during the immunization campaigns, if there is an outbreak like cholera, if there is an IDPs in a site and they need to have a hygiene awareness […*..*], but if we are talking about the role of other ministries, we are talking with endowment offices, to support us through Imam, Sheikh, and this also through C4D and other sectors*” **Informant # 7 (UN agency)**.

### 3. The coherence between agendas

In Yemen, there are several examples to clarify the interrelationship between the different health agendas: for example, literatures from Yemen show that the establishment of the electronic Disease Early Warning System (eDEWS) was established to strengthen health security in the country and focused on specific epidemic prone diseases by using representative samples of health facilities (sentinel sites) ^15^. The program was designed by the World Health Organization to support the routine surveillance system in Yemen, but due to the weakness of the disease surveillance system, eDEWS became the main source of information in the health system and currently covers most of the operating health facilities and including all diseases reported in the routine disease surveillance system, it called now the electronic Integrated Disease Early Warning System (eIDEWS) ^16^. Table 1 shows the increasing number of the involved governorates, districts and health facilities from 2013 to 2017, accordingly the number of consultations increased from one million in 2013 to cover more than 12 million in 2017.

**Table 1:** Expansion of the electronic Disease Early Warning System 2013-2017 ^16^

| Indicators | Dec 2013 | Dec 2014 | Dec 2015 | Dec 2016 | Dec 2017 |
| --- | --- | --- | --- | --- | --- |
| Number of governorates | 10 | 10 | 16 | 23 | 23 |
| Number of districts | 31 | 124 | 209 | 306 | 333 |
| Number of health facilities | 247 | 249 | 402 | 1406 | 1982 |
| Number of weekly bulletins | 41* | 51 | 51 | 49 | 49 |
| Number of reported events | 18 | 18 | 18 | 28 | 31 |
| Number of consultancies | 1,028,686 | 3,324,105 | 3,178,795 | 6,327,905 | 12,908,255 |

\*in 2013 system report started from week 11

Another example shows the need for interconnection between agendas. During the cholera epidemic, many donors pumped money to fight the epidemic that spread rapidly in Yemen. The implementing partners in the field found that focusing on the activities to combat the epidemic is not sufficient in light of the current situation that many people do not have access to health services, so the need has emerged to adopt a strategy to enhance access to health services everywhere in Yemen^17^. Then, we have seen a great support to the Minimum Health Services Package that developed by WHO and adopted by all the UN organizations operating in Yemen. Thus, an activity that started as a health security action to control outbreaks such as cholera, developed into a program in support of UHC though the implementation of the Minimum Health Services Package that developed by WHO and adopted by all the UN organizations operating in Yemen.

The catastrophic impact of the outbreak on health system and population in Yemen was highly expected during the pandemic of COVID-19. The health security in Yemen was ignored even before the war and preparedness for new emerging outbreak was not in place to protect the population^18^. Therefore, the spread of the outbreak has clearly affected the provision of health services in the country, many health facilities were closed in front of people, particularly when mortality increased among health workers ^19^. Additionally, lack of humanitarian funds is leading to shut down lifesaving interventions in Yemen, and jeopardize the universal health coverage^20^. Nevertheless, health promotion intervention played an important role in prevention and control under the absence of the essential health services ^21^.

> “*There was panic and Unpreparedness, Diversion of all efforts toward COVID-19 without effective plan for service continuity*… *Now I see there is less heat on hospitals, and most health facilities are providing services*.” **Informant # 8 (UN agency)**
>
> *“Health facilities suffered from a scarcity of medicines, solutions and supplies, with the high cost of transportation to reach these facilities because of the high prices of oil derivatives, which hindered many cases from reaching the health facility to seek health care consultations*.*”* **Informant # 10 (UN agencies)**
>
> *“Our aim was to raise awareness in the community about precautionary measures and ways to prevent transmission of infection in various places such as Qat markets, halls, schools, universities, mosques, and gathering places in general […*.*]. This strategy was also based on the involvement of multiple sectors, such as the Ministry of Media, the Local Administration, Ministry of Education, and Ministry of Transport. Different tasks were set for each one with the aim of delivering an educational awareness message to the population*.*”* **Informant # 11 (MoPHP)**

## Discussion

This paper focuses on the impact of fragmented health system on the implementation of health policies with respect to UHC, GHS, and HP. It aims to develop an overall understanding of the role of the key stakeholders in implementing the three agendas and to analyse the implementation of agendas. This is the first study in Yemen that assesses existing policies, gaps, challenges, synergies and fragmentation in implementing health policies, the key findings are the following:

1. There are many stakeholders working together in Yemen for providing humanitarian aids and to achieving their agendas. There are few differences in stakeholders between north and south of Yemen.
2. Generally, MoPHP and WHO are the main implementers of the three agendas in the country. UHC is the most adapted agenda by all national health policies, while HP is not a priority for health authorities in the country particularly during the current conflict, however, it become a priority during the COVID-19 outbreak.
3. Implementing the global health agendas are facing several challenges, in addition to the conflict situation and outbreaks, there are many other factors such as political interference, week leadership, lack of coordination, absence of advocacy, and financial hardship.

Most of the key informants noticed that there is no link or connection between the three global agendas at the national level. This is because health security was isolated from the UHC, and HP is always ignored by stakeholders, thus, all the available strategies and interventions were driven and controlled by international donors rather than the MoPHP. Based on the current situation in Yemen, there are several stakeholders working together in Yemen to provide humanitarian aid and to achieve their agendas. Donors are engaged in humanitarian action due to the lack of national capacities during the fragile situation ^22^. There are 254 humanitarian partners working in the humanitarian field during the current conflict in Yemen ^3^. Some studies show that local NGOs have easy and rapid access to the local communities and are the first to react to any emergency which is cost-effective in comparison to the international NGOs ^23,24^. It is a big humanitarian community compared to the humanitarian community in Iraq and Syria with 94 and 161 humanitarian partners respectively ^25,26^.

A lack of coordination between different donors and actors, and the absence of advocacy on the global agendas lead to fragmented leadership and governance and poor accountability within MoPHP. The presence of a huge number of donors and health actors may be considered as an asset to advocate for the humanitarian needs in the country and raise funds. There are several studies reflecting the role of the international community in supporting the fragile state to be more engaged in strengthening the health systems ^27^. However, this is depending on the coordination of the donors and the full understanding of the challenges in the fragile health system and it will help to strengthen the civil society institutions and to re-establish the legitimacy to governments ^13,28^. For example, the Afghani Ministry of Health used to coordinate with the international community to improve the health system in the country ^13^. The cooperation and communication pathway between the Ministry of Defence (MoD) and the MoPHP in Yemen is very weak to apply and implement the national health strategies (NHS) at multi-sectoral levels. The role of the MoD should be effective particularly in health security, for example, the MoD in Saudi Arabia is supporting the health system to scale up the standards of health care in the hospitals by applying for the quality management programs in the hospitals ^29^.

The NHS 2010-2025 is the main strategy for the MoPHP in Yemen, and it concentrates on increasing the access to health services and health security, but they ignore the HP part ^30^. The UHC principles are to increase access to good quality basic health services in all communities through all national health policies in Yemen, like the NHS and the Minimum Service Package (MSP). On the other hand, the UHC as a term is not used in the old and new national health policies. For example, the MSP that was modified in 2017 during the war from the Essential Service Package which developed earlier in 2000, therefore MSP did not mention the term of UHC, however, it has all the principles and concepts of UHC ^31^. There are some reasons for this such as the following: first, the concept of the UHC came with the SDGs in 2015 after the development of the NHS at 2010 in Yemen, second, lack of awareness and knowledge about these global strategies, and the inability and failure of the stakeholders to reflect the global agendas in the national level during the conflict. A lesson learnt should be taken from Myanmar which successfully implemented UHC in its’ new National Health Plan 2017 – 2021 ^32^.

The GHS principles are to protect the population from the health emergencies or the outbreaks in fragile health systems during conflicts or natural disasters that can cross the borders and can lead to public health emergencies of international concern that threats all the world ^33,34^. Furthermore, the health security agendas in the NHS were not prepared for the conflicts and outbreaks, and lack of support of the central laboratories in the country, which is considered as one of the gaps in this NHS for Yemen. This gap is obvious during the current COVID 19 outbreak in Yemen, there is severe shortage in lab capacities to confirm cases early in country ^35^. Some studies have shown that good coordination between laboratories and health information systems increase the ability of the countries to detect, report and respond to any threats that can affect the GHS ^35,36^. Despite the fragmentation of implementing the health agendas, the actors in Yemen released that controlling outbreaks such cholera cannot be achieved by supporting health security alone. Yemen received generous support to function health facilities and resume basic health services through backup the newly introduced Minimum Services Package ^37,38^.

The MoPHP and WHO are supposed to be the main supporters of implementing all the global policies related to the UHC, GHS and HP agendas on the national level. However, in reality, there is a gap in implementing those global agendas in national health policy by these two actors. Most of the health stakeholders are focusing on the UHC, HS, while the HP is not a priority for them in the current situation, the cholera outbreak is a good example to explain this issue. The international humanitarian organizations are good in the detection, response, and management of the cholera cases, while the gap is in the HP and prevention component to control the outbreak ^39^.

## Conclusions

Restoring peace, prioritizing global health strategies, coordination, collaboration between donors and health actors, and control over and monitoring of stakeholders are highly recommended. A multi-disciplinary approach must be pursued and implemented by stakeholders in order to effectively and efficiently improve access to basic health care particularly with the current COVID 19 outbreak in Yemen.

## Data Availability

Data available on request

## Acknowledgments

We express our gratitude to all who accepted to be as part of this research as interviewers and provided us with informative documents.

## Competing Interests

The authors declare that they have no competing interests.

## Funding

None declared

## Contributors

T.H and F. D, contributed in all the research process, particularly study design, data collection, data analysis and writing the first of this manuscript draft. R. A, and H. N contributed in data collection and revising the manuscripts from their experiences in Yemen, L. H, revised the manuscript and editing the English language. A.J, is the overall supervisor and monitor all steps of this research.

## Patient and public involvement

Patients and/or the public were not involved in the design, conduct, reporting or dissemination plans of this research.

## Notes

### Competing Interest Statement

The authors have declared no competing interest.

### Funding Statement

no external funding was received

